# Synthetic electronic health records generated with variational graph autoencoders

**DOI:** 10.1101/2022.10.17.22281145

**Authors:** Giannis Nikolentzos, Michalis Vazirgiannis, Christos Xypolopoulos, Markus Lingman, Erik G. Brandt

## Abstract

Data-driven medical care delivery must always respect patient privacy – a requirement that is not easily met. This issue have impeded improvements to healthcare software and have delayed the long-predicted prevalence of artificial intelligence in healthcare. Until now, it has been very difficult to share data between healthcare organizations, resulting in poor statistical models due to unrepresentative patient cohorts. Synthetic data, i. e., artificial but realistic electronic health records, could overcome the drought that is troubling the healthcare sector. Deep neural network architectures in particular have shown an incredible ability to learn from complex data sets, and generate large amounts of unseen data points with the same statistical properties as the training data. Here, we present a generative neural network model that can create synthetic health records with realistic timelines. These clinical trajectories are generated on a per-patient basis and are represented as linear-sequence graphs of clinical events over time. We use a variational graph autoencoder (VGAE) to generate synthetic samples from real-world electronic health records. Our approach generates health records not seen in the training data. We show that these artificial patient trajectories are realistic and preserve patient privacy, and can therefore support safe sharing of data across organizations.

## 1 Introduction

Access to real-world health data is often restricted by privacy-protecting regulations like Health Insurance Portability and Accountability Act (HIPAA) and General Data Protection Regulation (GDPR), but also due to technical limitations or simply lacking incentives for data-sharing. Even when pseudo-anonymized (by leaving out personal identifiers such as social security numbers, residence, age, etc), a malicious agent with sufficient knowledge could re-identify patients by connecting patient attributes, conditions, medical prescriptions etc for an individual. Techniques like federated learning [1], differential privacy [2] and homo-morphic encryption [3] are actively researched to overcome these barriers.

Carefully created synthetic data could reduce data scarcity by exempting data from privacy preserving regulations. By design, synthetic data mimics real data but is decoupled from real individuals and can be safely shared among healthcare providers, academics, and private stakeholders without leaking sensitive or personally identifiable information. High-quality synthetic data enables exploration and hypothesis generation, but could also be used to pre-train AI models and thus decrease the need for vast amounts of original data. A synthetic data set that mirrors the original data well could also help focusing efforts on more probable hypotheses before seeking confirmation in the source data. Therefore, synthetic data could meet both privacy concerns and re-balance the effort of data access in relation to the chance of relevant findings, and also explore data patterns before investing too much in new research routes.

Healthcare data sets are complex in both space (heterogeneous and strongly connected) and time (cause and effect of symptoms, diagnoses, medications, etc.). Understanding the relationships between the different parts of information about a patient created along a patient trajectory is essential in clinical medicine. It is relatively straightforward to mimic the static properties of a given data distribution, but far more difficult to mimic diverse and coupled time-series with non-equidistant time steps [4, 5]. To the best of our knowledge, this remains to be done in the context of electronic health records (EHRs).

Deep learning (DL) models have revolutionized a wide range of real-world applications, from autonomous vehicles [6], to machine translation [7], and molecule generation in drug discovery [8]. Nevertheless, even the most successful DL model is at the mercy of the amount and quality of its training data set. DL algorithms are very data hungry, and the training samples must adequately reflect the full population that is to be learned. When such conditions can be met, DL algorithms have a canning ability to capture complex data patterns and they also generalize well to unseen data. In practice, available data is often insufficient to train DL models with millions of parameters in any meaningful way. As a consequence, DL models are especially sensitive to limited data availability, as manifested in healthcare.

Machine learning algorithms have already been successfully introduced in the healthcare informatics domain [9– 15]. Variational Autoencoders (VAEs) [16, 17] – and their graph off-springs [18–20] – and Generative Adversarial Networks (GANs) [21, 22] are recent deep learning architectures of particular promise. These models learn a “hidden”, underlying, data distribution from the training data. VAEs consist of an encoder-decoder pair. The encoder maps the input data to a latent (hidden) distribution, which is randomly sampled by the decoder with the objective to reconstruct the original input data. The latent distribution is usually chosen as a multivariate normal distribution characterized by its mean value and standard deviation. Once the model is trained, an arbitrary number of new samples can be generated by feeding the decoder random samples from the normal distribution. GANs, on the other hand, use two neural networks that are trained together but in adverse. The two networks are known as the generator and the discriminator. The generator learns to create samples as realistic as possible, while the discriminator learns to distinguish synthetic samples from real ones. Once both networks are fully trained, the generator can create unseen data samples with a high similarity to the real data.

Earlier efforts to simulate EHR data have used Bayesian network learning [23], and deterministic differential modeling, e. g., as implemented in the popular open-source software Synthea [24]. This open-source software package is designed to simulate the lifespans of synthetic patients but is based on fixed demographic properties extracted from public data, and does not learn through a training procedure. GANs have so far been the primary deep learning method to generate synthetic EHRs [9, 25–27]. Notably, Choi et al. proposed medGAN [9], a neural network model that generates high-dimensional discrete variables to represent EHR events. Baowaly et al. [25] derived two enhanced versions of medGAN with a more complex (Wasserstein) architecture, and Yale et al. [26] identified limitations to medGAN and proposed HealthGAN, another Wasserstein-based method. They also developed improved metrics for synthetic health data quality. Chin-Cheong et al. [28] created synthetic EHR data with GANs trained on patient data from intensive care units. The final results were combined with federated learning [1] to mimic a real-world scenario with data sets from different organizations isolated in silos. Finally, Esteban et al. [11] generated synthetic medical time series with a GAN using recurrent neural networks for both the generator and the discriminator. While GANs have achieved promising results, they also have drawbacks. GAN-generated data is continuous by default and must be paired to an autoencoder [9] or LSTM generator to produce discrete results [11]. Also, the adversial style used to train GANs increases the risk of getting stuck in local minima during learning [29]. Such minima are very challenging to escape by more training alone, and may limit the use of GANs on certain data structures (such as graphs).

Interestingly, Variational Graph Autoencoders (VGAEs) have not yet been used to generate synthetic EHRs. VGAEs present a promising route in this area, because they are easy to train, have been applied successfully to other graph learning problems, and can accurately model the underlying data distribution. We discuss the strengths and weaknesses of GANs vs VGAEs in more detail in the Results section.

In this paper, we develop a machine learning algorithm for generating electronic healthcare records represented as sequential graphs (patient trajectories). A patient trajectory is a time sequence of encounters (visits) at healthcare organizations (e. g., hospitals or other providers). Each encounter links to patient interventions such as identified medical conditions and requested medications. Analyzing such patient trajectories is key to deliver data-driven insights to healthcare organizations. Creating synthetic EHRs with graph deep learning is to the best of our knowledge a new concept. Synthetic graphs are already trending in drug design [30–33], but patient trajectories require much larger (e. g., hundreds of nodes) graph representations than their drug molecule counterparts. This poses a significant challenge to generation algorithms. Here, we propose a VGAE tailored to patient trajectories that can generate novel large-scale samples.

## 2 Results

### 2.1 EHR data source

#### Details

The Medical Information Mart for Intensive Care (MIMIC-IV) database was the source to all our numerical experiments [34]. MIMIC-IV provides critical care data for thousands of patients admitted to the intensive care units at the Beth Israel Deaconess Medical Center. The patient, visit (encounter), diagnosis (condition), and medication (medication request) data tables were migrated to a labeled property graph (LPG) database. These data tables cover healthcare events at the emergency unit and the following in-patient stays. They are a subset of a complete graph model for healthcare data developed by one of the authors [35] which follows the FHIR standard for healthcare data [36]. The LPG database was constructed by connecting all visits (encounters) for a patient in chronological order. Conditions and medications for the patient are then attached to the encounters. Descriptive nodes for the types of conditions and medications are shared among the patients in the database. Labels are the only node and edge attributes included in this work. Our results are easily extended to include more metadata stored as key-value pairs on nodes and edges.

A patient graph (trajectory) is defined as the subgraph constructed with its origin at a patient node, following edges outwards until reaching terminal nodes (type-describing nodes which have no outgoing edges). Patient graphs defined in this way are directed acyclic graphs (DAGs), i. e., they do not contain directed cycles. We extracted a subset of patients whose trajectories contain any of the ICD-10-CM diagnosis codes I48.0, I48.1, I48.2, and I48.9. This group corresponds to a real-world cohort of patients diagnosed with atrial fibrillation.

A trajectory graph was constructed for each of those 6535 patients. The node and edge labels are described by the functions *ℓ*_*V*_ and *ℓ*_*E*_ which assign elements from the sets ∑_*V*_ and ∑_*E*_ (i. e., *ℓ*_*V*_ : *V*→ ∑_*V*_ and *ℓ*_*E*_ : *E* →∑_*E*_). Here, *V* and *E* denote the set of nodes and edges of all graphs, respectively. There are in total |∑_*V*_| = 13, 980 different node labels and |∑_*E*_| = 6 different edge labels. Figure 1 shows an example of a patient trajectory. Each trajectory contains one Patient node, and a number of Encounter nodes that form the patient timeline. Each Encounter is described by an EncounterCategory and is shaped like a star graph with Condition/ConditionType and MedicationRequests/MedicationType pairs for the diagnosis and medication events. The edge labels are ATTENDS between Patient and each Encounter, NEXT between neighboring Encounters, OF CATEGORY to describe the Encounter, DIAGNOS between Encounter and Condition, ADMINISTRATED between Encounter and Medication, and OF TYPE to describe the Condition/ConditionType and MedicationRequests/MedicationType pairs. The edge labels are uniquely determined by the label pair of the ancestor and successor nodes. Note that edge labels are omitted from Figure 1 to simplify the presentation. Figure 2 shows the frequency of the node and edge labels in the training data (see Table 1 for more details on the source data). ConditionType and MedicationType labels are higher level labels that contain all diagnosis and medication events, respectively (i. e., labels I10, E92, Heparin, Mupirocin Ointment 2%, etc. in Figure 1).

**Figure 1:**
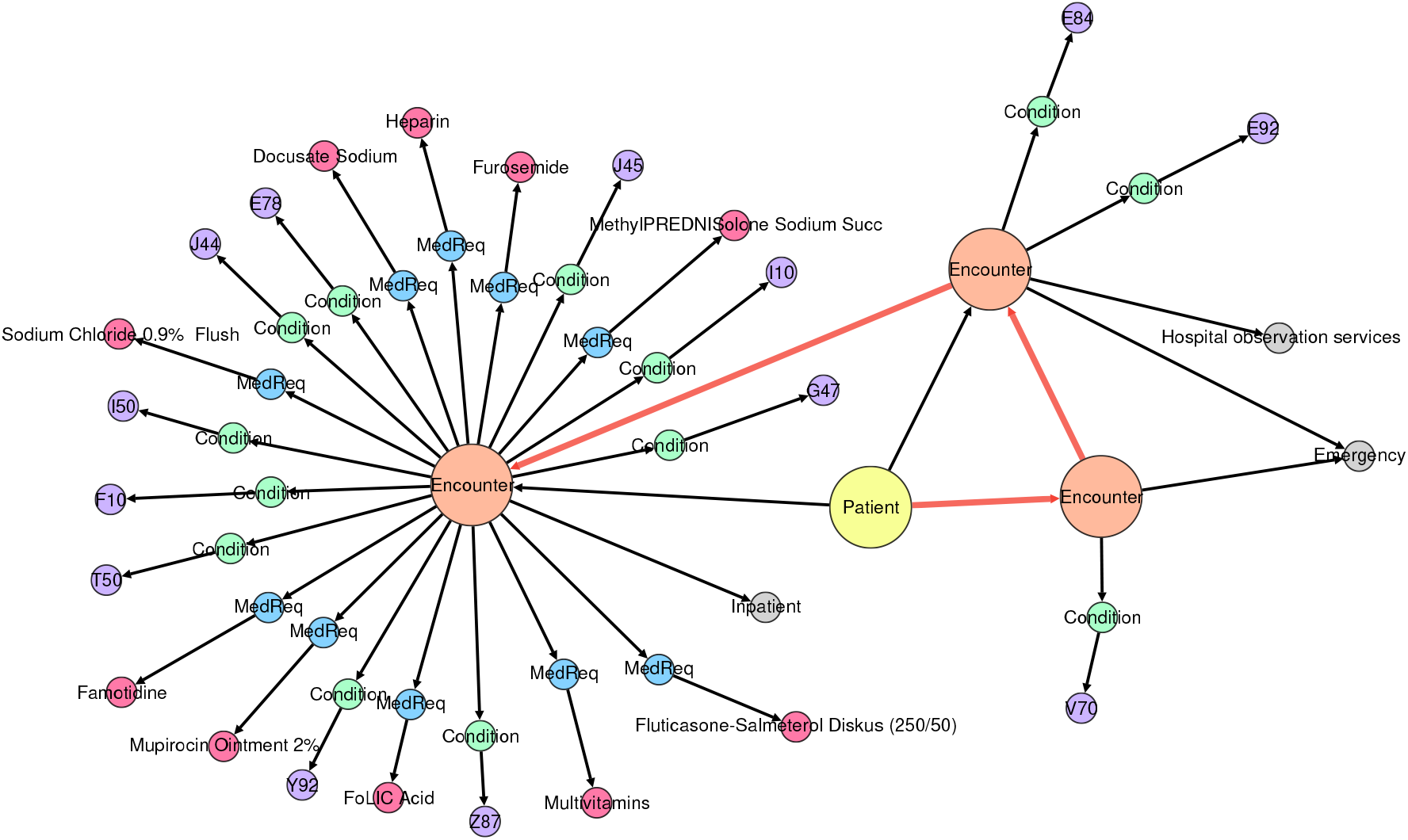
Visualization of a patient trajectory. Each trajectory is represented as a directed acyclic graph. The patient timeline is represented by the edges between Encounters (highlighted in red).

**Figure 2:**
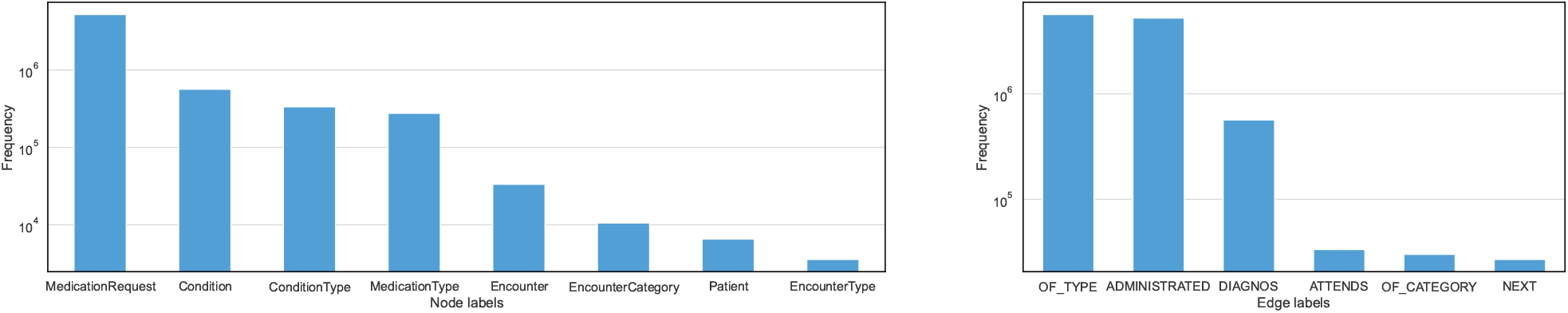
Histogram of the number of (higher level) node labels and edge labels in the training data. There are eight different (higher level) node labels and six different edge labels in total. Note that some higher level node labels such as MedicationType, ConditionType aggregate hundreds or even thousands of lower level node labels.

**Table 1:** Statistics on the patient trajectories calculated from the atrial fibrillation cohort extracted from the MIMIC-IV database. Statistics are provided for both raw data and processed data.

|  | Raw | After<br>Pre-processing |
| --- | --- | --- |
| Max # nodes | 18,947 | 2,772 |
| Min # nodes | 10 | 10 |
| Average # nodes | 1,044.1 | 221.2 |
| Max # edges | 36,811 | 5,162 |
| Min # edges | 9 | 9 |
| Average # edges | 1,867.3 | 294.7 |
| # node labels ($\vert \Sigma_V\vert $) | 13,980 | 944 |
| # edge labels ($\vert \Sigma_E\vert $) | 6 | 6 |
| # graphs | 6,535 | 6,535 |

#### Pre-processing

There is a large number of distinct ICD-10-CM diagnostic codes (and analog ATC-codes for medications) in the MIMIC-IV data. Since the atrial fibrillation cohort is limited to 6535 patients, pre-processing is needed to reduce the number of node labels for the learning algorithm. The data was processed by: (1) Dropping all Condition nodes which correspond to earlier versions than ICD-10-CM (ICD-9 and a few ICD-8 codes). (2) Only keeping the chapter (the three first characters) of the ICD-10-CM codes, and merging nodes that ended up as identical. (3) Dropping rare events (condition and medication nodes that occurred less than 50 times). After these steps, 944 node labels remained. The largest graph had 143 Encounters and the largest Encounter had 180 successors. More details about the pre-processed trajectories are given in Table 1.

### 2.2 Generating model

Graph learning algorithms are usually permutation invariant to the ordering of nodes. Since nodes are added one at a time, node ordering becomes important. By modeling patient graphs (patient trajectories) as DAGs, graph generation is significantly simplified, because every DAG has a at least one linear ordering of the nodes such that for every directed edge (*u, v*), node *u* comes before node *v*. This is known as a topological ordering and can be computed in linear time.

We found that standard recurrent neural networks were unable to learn realistic patient trajectories (see the discussion in the Methods section). We therefore designed a model tailored to the structure of our patient graphs. These trajectories are built up of linear sequences of Encounter nodes, where each Encounter node is the center of a star graph. These two substructures (linear sequence and star) can be modelled separately.

GANs and VAEs are two of the most common architectures for generative models. They both learn from training data and can generate new and previously unseen data samples. GANs can generate highly realistic images, audio, and text, but can be difficult to train, as the generator and discriminator networks may get stuck in a local equilibrium [29]. GANs struggle to provide meaningful latent data representations, while VAEs can interpolate smoothly between data points to find generate new representative patient trajectories. Importantly, GANs work best with continuous data. We formulate graph generation as a sequential task where nodes and edges are added to the graph in iterations. This is natural for DAGs but poses a challenge for backpropagation because the gradients are zero. The usual workarounds can not be applied to graphs (e. g., Gumbel softmax [37]). One option is to avoid sequential generation and generate the adjacency matrix in one shot [38]. However, this approach is only feasible for small-graph applications, e.g. molecule generation (the adjacency matrix is a (*n* × *n*)-matrix for a graph of *n* nodes).

In light of the above discussion, we used a variational autoencoder as the basis of our synthetic data generator. The encoder maps patient trajectories into a parameterized multivariate Gaussian distribution (i. e., the encoder predicts the mean vector and covariance matrix of this distribution). A random sample is drawn from the distribution (the “hidden” representation of the input patient trajectory) and fed into the decoder to reconstruct the original patient trajectory. Once trained, new trajectories can be generated at scale by drawing random samples from a parameterized normal distribution and using the decoder to output a synthetic trajectory. Further details on the model architecture and training details are found in the Methods section.

### 2.3 Experiments

#### Graph reconstruction

We first investigated whether the proposed model can accurately reconstruct its input graphs. To this end, we used graph kernels; a positive semi-definite kernel on the set of graphs 𝒢 [39]. Roughly speaking, a graph kernel measures the similarity of graphs. Once we define a function *k* : 𝒢 × 𝒢→ ℝ on the set 𝒢, there exists a map *ϕ* : 𝒢 →ℋ into a Hilbert space ℋ, such that *k* (*G, G*^′^) = ⟨*θ* (*G*), *θ* (*G*^′^)⟩ _ℋ_ for all *G, G*^′^ ∈ 𝒢 where ⟨· ·⟩_ℋ_, is the inner product in ℋ. Graph kernels are grouped into major families that focus on different structural aspects of graphs. We primarily relied on the Weisfeiler-Lehman subtree (WL) kernel [40] and on the shortest path (SP) kernel [41] to compare input graphs against reconstructed graphs. WL and SP are among the most successful graph kernels and account for both graph structure and node label information.

We computed the histogram of *k*_*i*_ = *k* (*G*_*i*_, *Ĝ* _*i*_), where *G*_*i*_ is an input graph, *Ĝ* _*i*_ its corresponding reconstructed graph, and *k*(·, ·) is a graph kernel (i. e., WL or SP) with *i* ∈ {1, 2, …, 6535}. Here, *k*_*i*_ = 0 means that reconstructed graph *i* is completely different to its input, and vice versa *k*_*i*_ = 1 implies identity up to isomorphism. For the reconstruction task, ideally, we would like the model to output graphs isomorphic to those given as input. Thus, we would like most kernel values to be large (close to 1). The histogram in Figure 3 shows a very high similarity (*k* > 0.9 for 3/4 of the graph distribution) for most graphs. This indicates that the proposed model yields very good performance in reconstructing the input graphs even when some of them are large and consist of many Encounter nodes.

**Figure 3:**
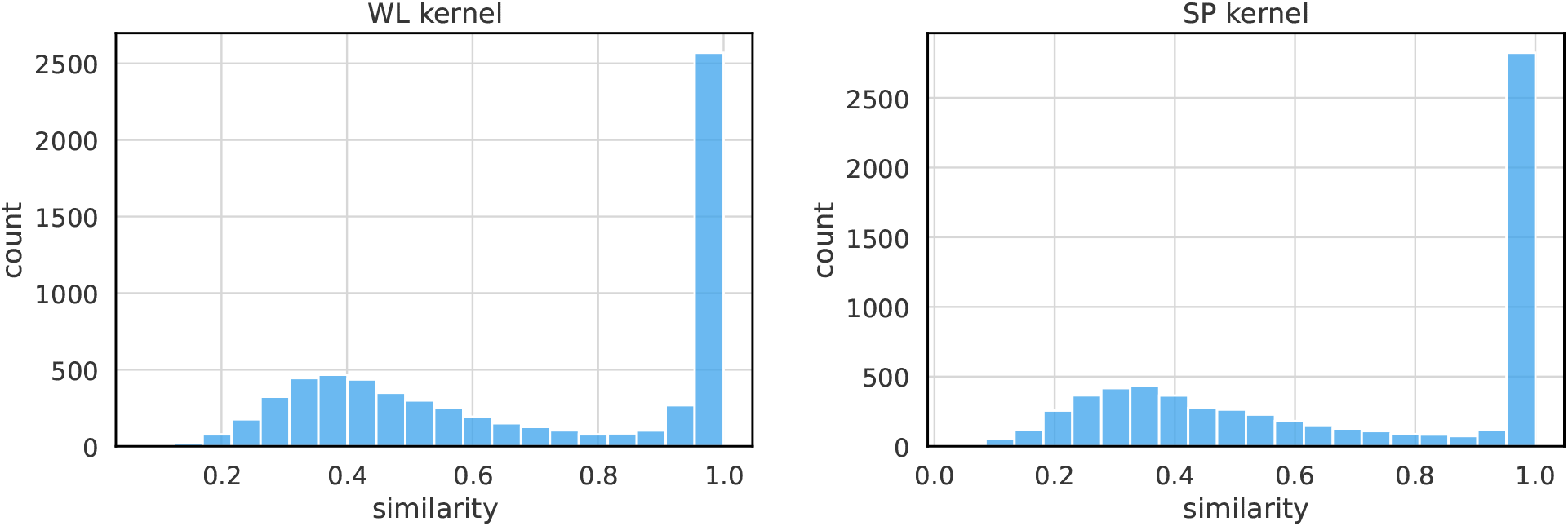
Histograms of similarities between input and reconstructed trajectories. Each input trajectory was compared against its corresponding reconstructed trajectory using a graph kernel. (a) Histogram of similarities between input graphs and reconstructed graphs using the Weisfeiler-Lehman subtree (WL) kernel. (b) Histogram of similarities between input graphs and reconstructed graphs using the shortest path (SP) kernel.

#### Graph generation

We have established that the model successfully reconstructs the patterns from the input graphs. Can the model also generate novel synthetic graphs that are realistic but not found in the training data? To investigate this, we generated 10000 synthetic patient graphs by feeding random samples drawn from the multivariate normal distribution to the decoder.

We used the WL and SP graph kernels to compare the generated synthetic graphs to the input graphs from the training data. We computed the histogram of the maximum similarity 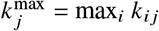 for the two kernels, where now *G*_*i*_ is an input graph, *Ĝ* _*j*_ is a graph generated from a random sample with *j* ∈ {1, 2, …, 10000}, and *k*_*i j*_ = *k* (*G*_*i*_, *Ĝ* _*j*_) is the (*i, j*):th element of a (6535 × 10000)-similarity matrix with *k*(·, ·) being a graph kernel. We end up with 10000 kernel values.

Figure 4 shows that both kernels’ maximum similarity distributions are centered around *k*^max^ ∼ 0.55. There is a small fraction of samples for which *k*^max^ ≈ 1 holds. Such graphs correspond to near-identical replicas of input graphs and could lead to patient privacy leaking from the training set. These graphs must be eliminated from the generated data set to reduce the risk of privacy leaking.

**Figure 4:**
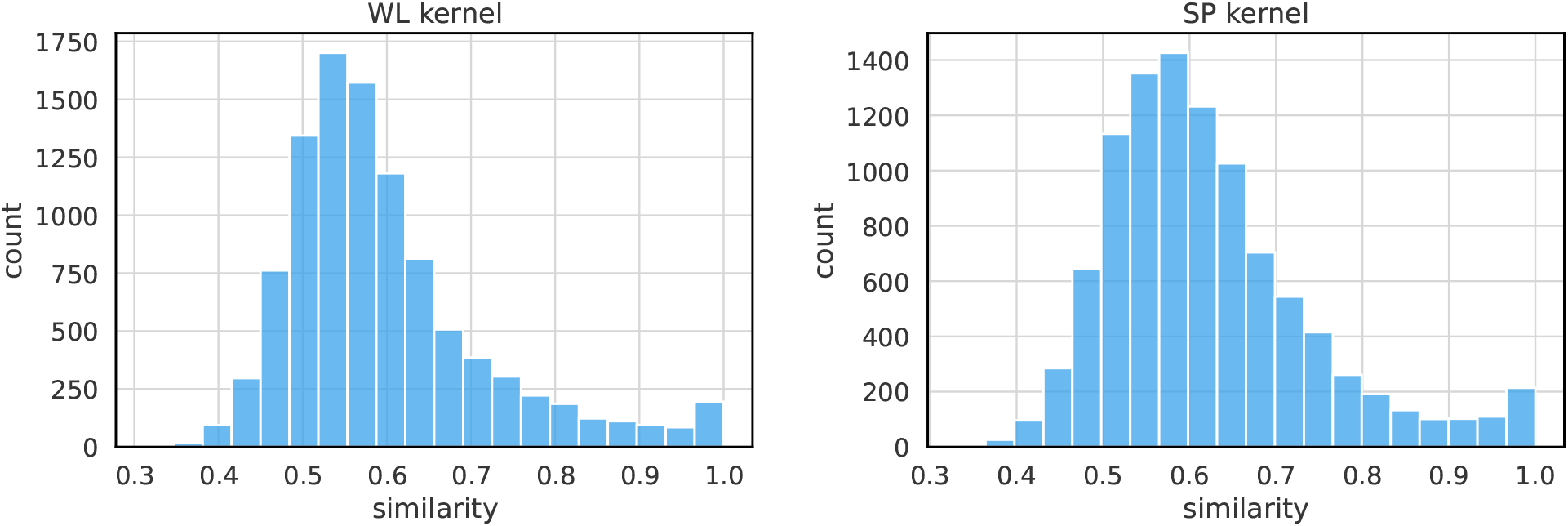
Histograms of similarities between input and generated trajectories. The model was used to generate 10000 synthetic trajectories, and for each synthetic trajectory, the most similar input trajectory was found. (a) Histograms of similarities between input and generated trajectories using the Weisfeiler-Lehman subtree (WL) kernel. (b) Histograms of similarities between input and generated trajectories using the shortest path (SP) kernel.

A simple method to avoid these replica graphs is to add Gaussian noise to the decoder-generated Encounter node representations. We can guarantee that all generated graphs are novel samples by tuning the variance of the Gaussian. Figure 5 shows the proportion of novel samples as a function of the standard deviation of the Gaussian distribution. We generated 20000 samples for each value of *σ*. As expected, the number of novel samples is an increasing function of *σ*, and all samples are novel when *σ* ≥ 0.5. More rigorous methods to ensure privacy could also be used. For example, there are VAE algorithms that perturb the latent vectors that represent the samples in a differential privacy-consistent way [42]. The fraction of replicas is small even without added noise. More than 99% of the generated samples are novel. In addition, Figure 4 shows that there are no graphs at low *k*^max^. Contributions near *k*^max^ = 0 would have indicated very low similarity to the inputs and generated patient trajectories that are unrealistic.

**Figure 5:**
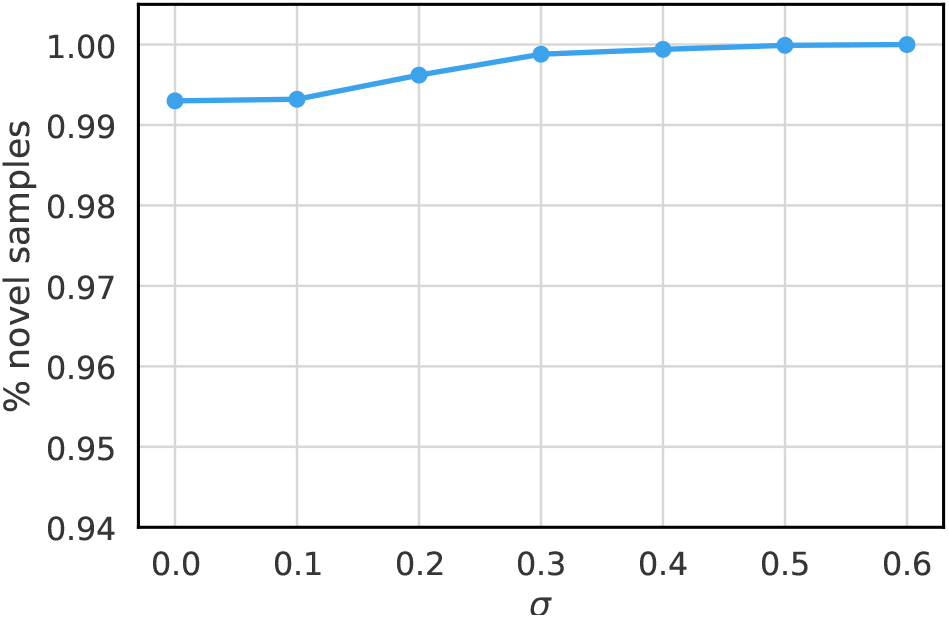
Percentage of novel trajectories as a function of the standard deviation of the Gaussian noise. The mean of the Gaussian was chosen to be the zero vector. The noise was added to the representations of the Encounter nodes. For each value of *σ* ∈ {0.0, 0.1, 0.2, 0.3, 0.4, 0.5, 0.6}, 20000 trajectories were generated in total which were compared against the real trajectories.

Further, we must determine to what extent these novel generated samples are realistic representations of electronic health records. In what follows, we remove samples that are very similar to input graphs (those graphs *Ĝ* _*j*_ for which 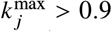 according to the WL or SP kernel). We use the Graph Kernel-Maximum Mean Discrepancy (GK-MMD) metric to evaluate the quality of the generated graphs. The GK-MMD accounts for both graph topology and node labels [43]. The MMD statistical test determines whether sample sets from two distributions *P* and *Q* were in fact derived from the same distribution (i. e., whether distributions *P* and *Q* are identical). The square of the MMD is:

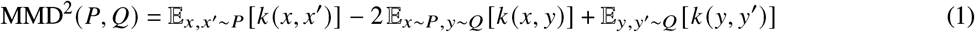

where *k*(·, ·) is the associated kernel function [44]. A lower MMD score indicates a better approximation in terms of the employed kernel (0 means that they are identical). We computed the squared WL-MMD and SP-MMD scores between the generated trajectories and their training data. The scores were 0.0017 and 0.0020, respectively.

We also investigate if paths of length *n* occurred with the same frequencies in the generated graphs as in the input graphs. Such *n*-paths can be thought of as Condition and Medication node pairs separated in time by (*n* − 1) Encounters. In this terminology, 1-paths correspond to node labels, 2-paths to nearest-neighbors, and 3-paths to nextnearest-neighbors. The first is a static (time-independent) property, but the other two are dynamic properties through the timeline implicit by the NEXT-relation between neighboring Encounters. A few examples of 2- and 3-paths are highlighted in Figure 6. The number of such paths increase exponentially which has a significant impact on the time complexity to compute correlation coefficients for larger *n*. We calculated Pearson’s *r* as a function of *n* (Table 2 and Figure 7). The static 1-paths (node labels) are perfectly retained in the generated graphs (*r* = 0.995). This shows that static properties like patient attributes, and number of diagnosis and medications are indistinguishable in the generated graphs compared to the training data. The dynamic 2- and 3-paths are almost equally well preserved in the novel training samples (*r* > 0.93). This shows without a doubt that the model learns time-dependencies between conditions and medications that occur in consecutive Encounters.

**Table 2:**
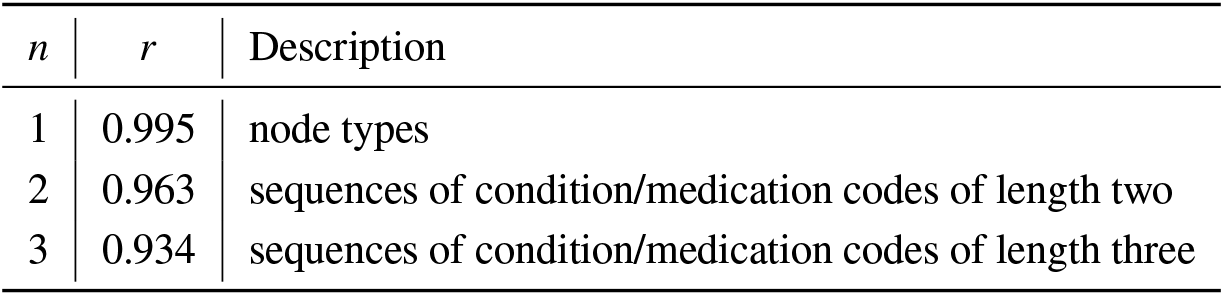
Pearson’s *r* between frequency of different structures in the collection of training trajectories and the collection of generated trajectories. Structures correspond to 1-paths (node types), 2-paths and 3-paths as illustrated in Figure 6.

| $n$ | $r$ | Description |
| --- | --- | --- |
| 1 | 0.995 | node types |
| 2 | 0.963 | sequences of condition/medication codes of length two |
| 3 | 0.934 | sequences of condition/medication codes of length three |

**Figure 6:**
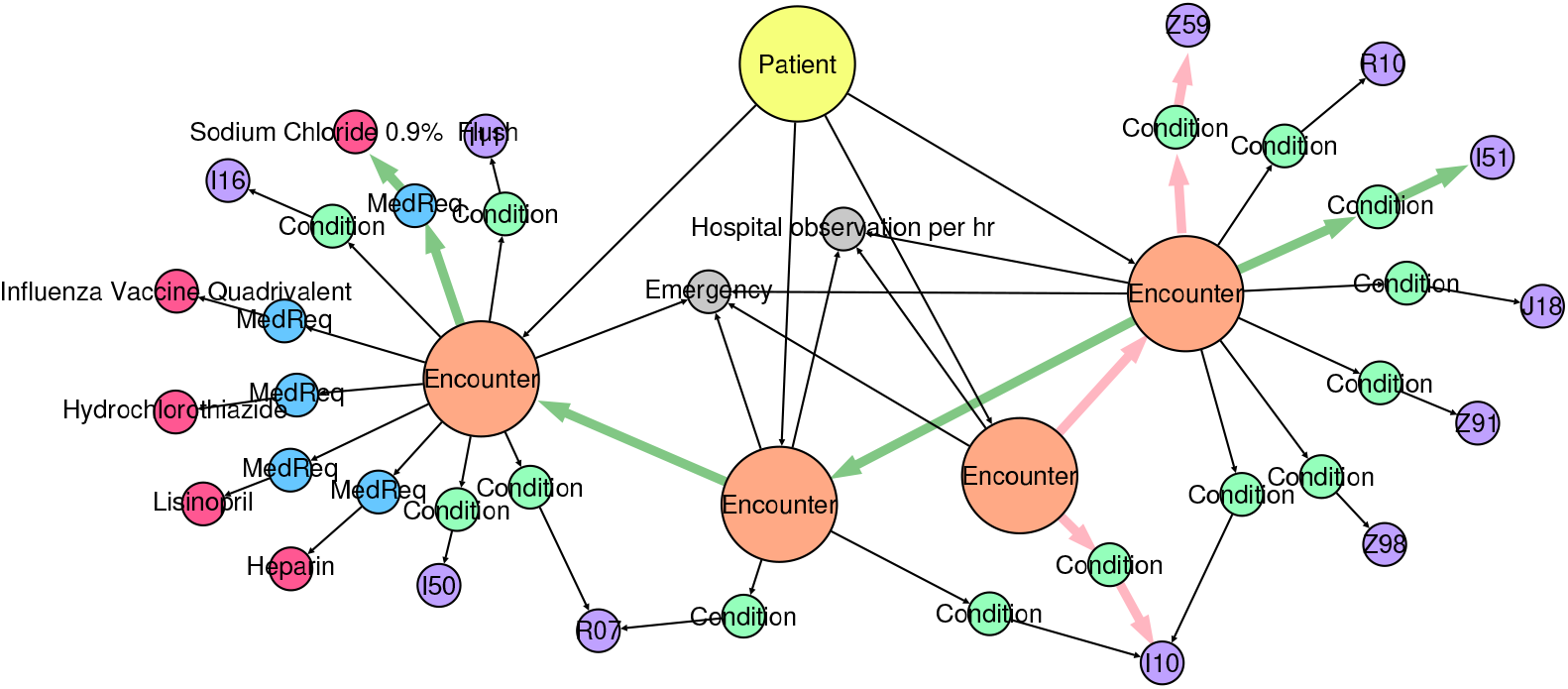
Examples of 2- and 3-paths in a patient trajectory. A 2-path from the ConditionType node I10 of the first Encounter node to ConditionType node Z59 of the second Encounter node is highlighted in pink. A 3-path from the ConditionType node I51 of the second Encounter node to MedicationType node Sodium Chloride 0.9% of the fourth Encounter node is highlighted in green.

**Figure 7:**
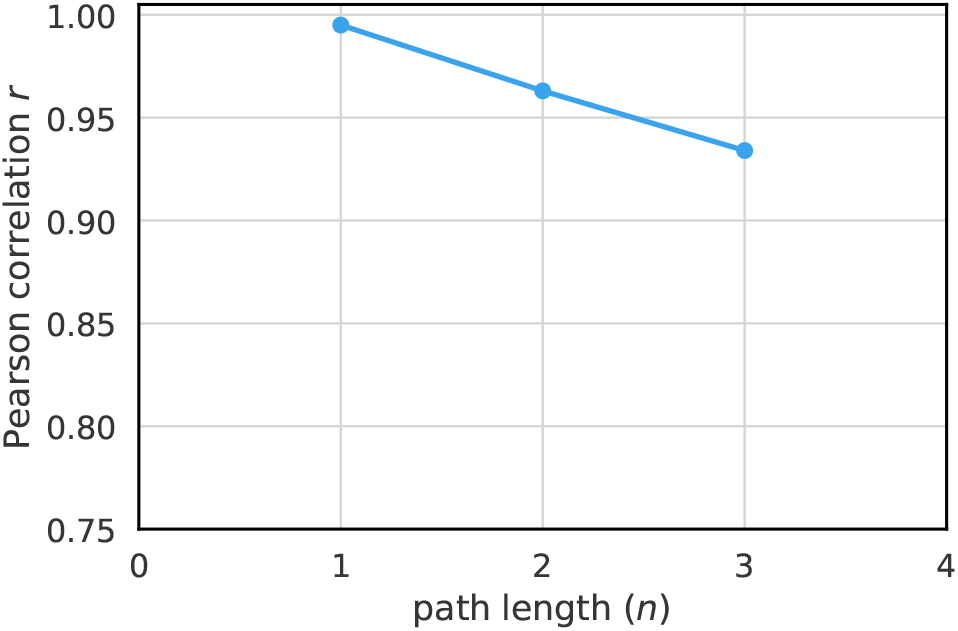
Pearson’s *r* between frequency of different structures in the collection of training trajectories and the collection of generated trajectories. Structures correspond to 1-paths (node labels), 2-paths and 3-paths as illustrated in Figure 6.

#### Clinical validation

We can show the generated data quality more concretely. First, the patient visit length is the number of consecutive Encounters in the patient’s trajectory. Figure 8 shows the frequency of visit lengths for real and synthetic patient cohorts, respectively. The two distributions are very similar, and both real and synthetic cohorts are dominated by short visit lengths (less than 20 Encounters, even though the input data contains a 144-length outlier). We computed a MMD of 0.025 between the two distributions. This very low value further verifies that the generated trajectories are realistic.

**Figure 8:**
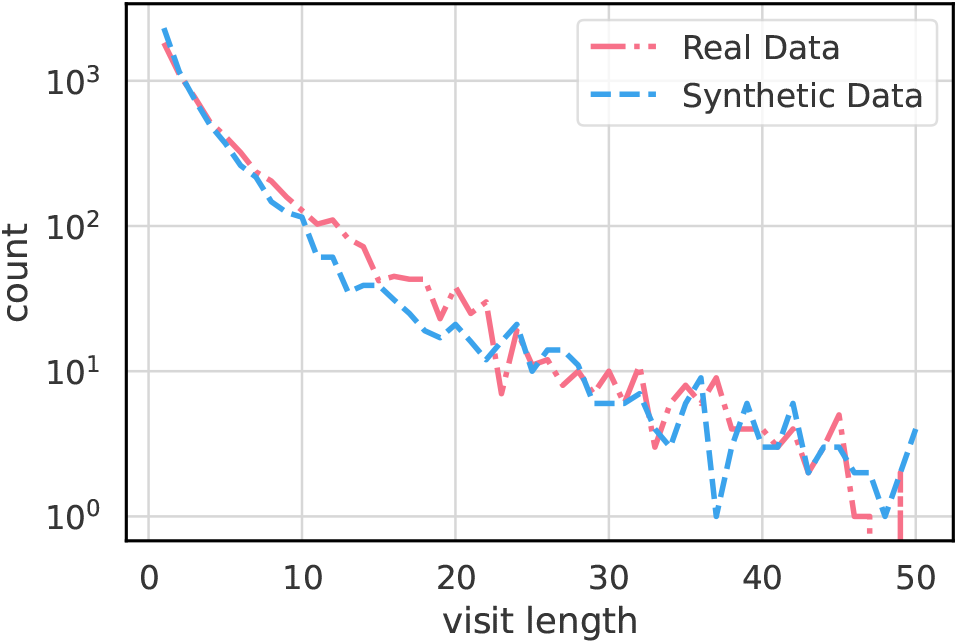
Plot of visit lengths for real and synthetic data. The visit length refers to the number of Encounter nodes that exist in a patient trajectory. Only trajectories consisting of at most 50 Encounter nodes are considered.

Next, we calculated comorbidities, defined as two or more medical conditions that exist simultaneously in the patient trajectory regardless of causal relationships. Following Ref. [45], we define the comorbidities for atrial fibrillation as Heart failure (ICD-10-CM code I50), Hypertension (I10-I15), Diabetes mellitus (E10-E14), and Stroke/TIA (G45, I63, and I74). Figure 9 shows comorbidities for the real and synthetic atrial fibrillation cohorts. The synthetic data faithfully replicates the trends in the original data. The only difference is that patients with combined heart failure and hypertension are slightly more common with diabetes in the real data than in the synthetic data.

**Figure 9:**
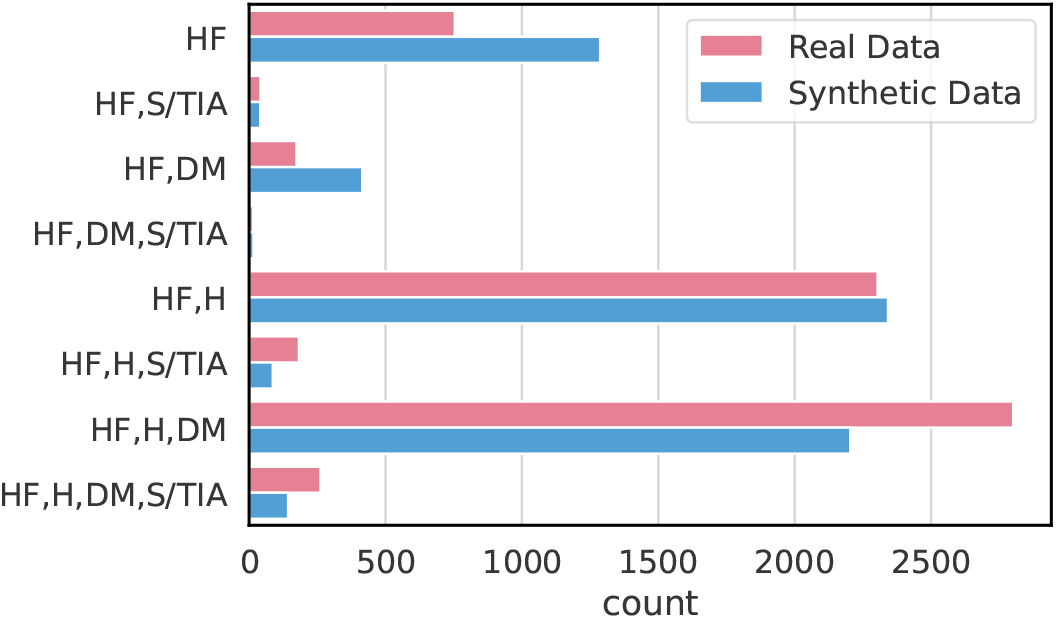
Comorbidities for the real and synthetic atrial fibrillation cohorts. We define the comorbidities for atrial fibrillation as follows. *HF*: heart failure, S*/TIA*: Stroke/TIA, *DM*: Diabetes mellitus, *H*: Hypertension.

#### Downstream analytics

We now investigate whether synthetic data samples can stand in for real-world data in downstream analytics tasks. As mentioned, heart failure is a common and potentially fatal comorbidity to atrial fibrillation. It is the leading cause of hospitalization and readmission in older adults [46]. In fact, about 40 million people worldwide suffered from heart failure in 2015 [47]. Machine learning methods have already improved predictions of hospital readmissions in heart failure patients [48], but onset of heart failure is more difficult to predict.

Here, we designed a classification task where the objective is to predict whether heart failure is the cause of the next patient encounter. We developed a classifier by pairing the encoder module of our VGAE with a multi-layer perceptron (MLP). This model was trained from scratch to make predictions in two different scenarios: “train on real, test on real”, and “train on synthetic, test on real”. These scenarios demonstrate whether a model trained on synthetic data generalizes to real data with similar performance to a model trained on real data. We used identical test sets in the two scenarios, and the same number of training samples. All data sets were balanced with equal number of input samples from each class.

For each scenario, we designed two experiments. In Task 1, the last Encounter node was removed from each trajectory. The sample was assigned to class 1 if that Encounter node linked to a Condition node with ICD-10-CM code I50 (heart failure), and to class 0 otherwise. In Task 2, we removed all trajectories with either no I50 code, or where the I50 code was connected to the first or second Encounter. By definition, the first I50 code of the trajectory is connected to the *i*:th encounter (*i* > 2). The trajectories were then assigned to class 0 (heart failure did not occur) or class 1 (heart failure did occur) with equal probability. For each class 0-trajectory, we generated a random number *k* = {1, …, *i* − 2}. The trajectory’s first *k* Encounters and their Conditions and MedicationRequests were kept. For each class 1-trajectory, the first *i* − 1 Encounters and their Conditions and MedicationRequests were kept.

We trained the two tasks as binary classifications. Training and analysis were repeated 10 times. The average test accuracy and standard deviation is reported in Table 3. We find that the classifier’s performance is independent of whether real or synthetic training data is used. In both tasks, training on synthetic data do not impact the performance of the classifier. Thus, for these downstream tasks, the synthetic data provides the same predictive performance as the real data. The data patterns needed for these predictions are learned by the synthetic generator.

**Table 3:** Classification accuracy of the two experimental scenarios in two separate downstream analytics tasks. Both tasks are variants on predicting the onset of heart failure in patients. In the first scenario, the classifier is trained on real data and is evaluated on real data, while in the second scenario, the classifier is trained on synthetic data and is evaluated on real data

| Scenario | Task 1 | Task 2 |
| --- | --- | --- |
| Train on real, test on real | 64.02% ± 2.93 | 73.04% ± 1.35 |
| Train on synthetic, test on real | 63.85% ± 2.57 | 73.32% ± 2.90 |

#### Distinguishing synthetic data from real data

Finally, we validated our model with outlier identification using a one-class Support Vector Machine (SVM), which is an unsupervised variant of the standard SVM [49]. SVMs can be applied to graph data using graph kernels. We generated 6535 synthetic patient trajectories and checked that no samples were isomorphic to the real training set. We randomly selected 10% of the synthetic trajectories and added them to the real trajectories (this is the “inlier” set). Then, we sampled 10% of the remaining synthetic trajectories and used the trained one-class SVM on each candidate to decide whether it is an inlier or not (“outlier”).

We used the SVM parameter *v* = 0.1 meaning that no more than 10% of the training samples can be considered as outliers by the decision boundary. We repeated the prediction experiment 10 times by sampling different subsets of synthetic trajectories. For comparison, we split the real data into training and test sets at an 90:10 ratio. As before, we enriched the training set with 10% of the synthetic trajectories and let the SVM classifier identify outliers in real data. Table 4 shows the results of these numerical experiments. The one-class SVM algorithm predicts that a large majority (92-93%) of the synthetic trajectories are inliers, independent of graph kernel. The algorithm performance is similar (90-91%) on the real data samples. This shows that the generated trajectories are of high quality and indistinguishable from real data. Figure 10 shows examples of how similar trajectories from the original data set are to novel samples generated with our VGAE-based model.

**Table 4:** Percentage of real and synthetic samples that were identified as outliers by the one-class SVM algorithm. Two graph kernels are employed to compute the kernel values (i. e., similarities) between trajectories, namely the Weisfeiler-Lehman subtree (WL) kernel and the shortest path (SP) kernel.

| Kernel | Outliers |  |
| --- | --- | --- |
|  | Synthetic Data | Real Data |
| WL | 7.8% ± 1.2 | 10.7% ± 0.9 |
| SP | 6.9% ± 1.0 | 11.3% ± 0.8 |

**Figure 10:**
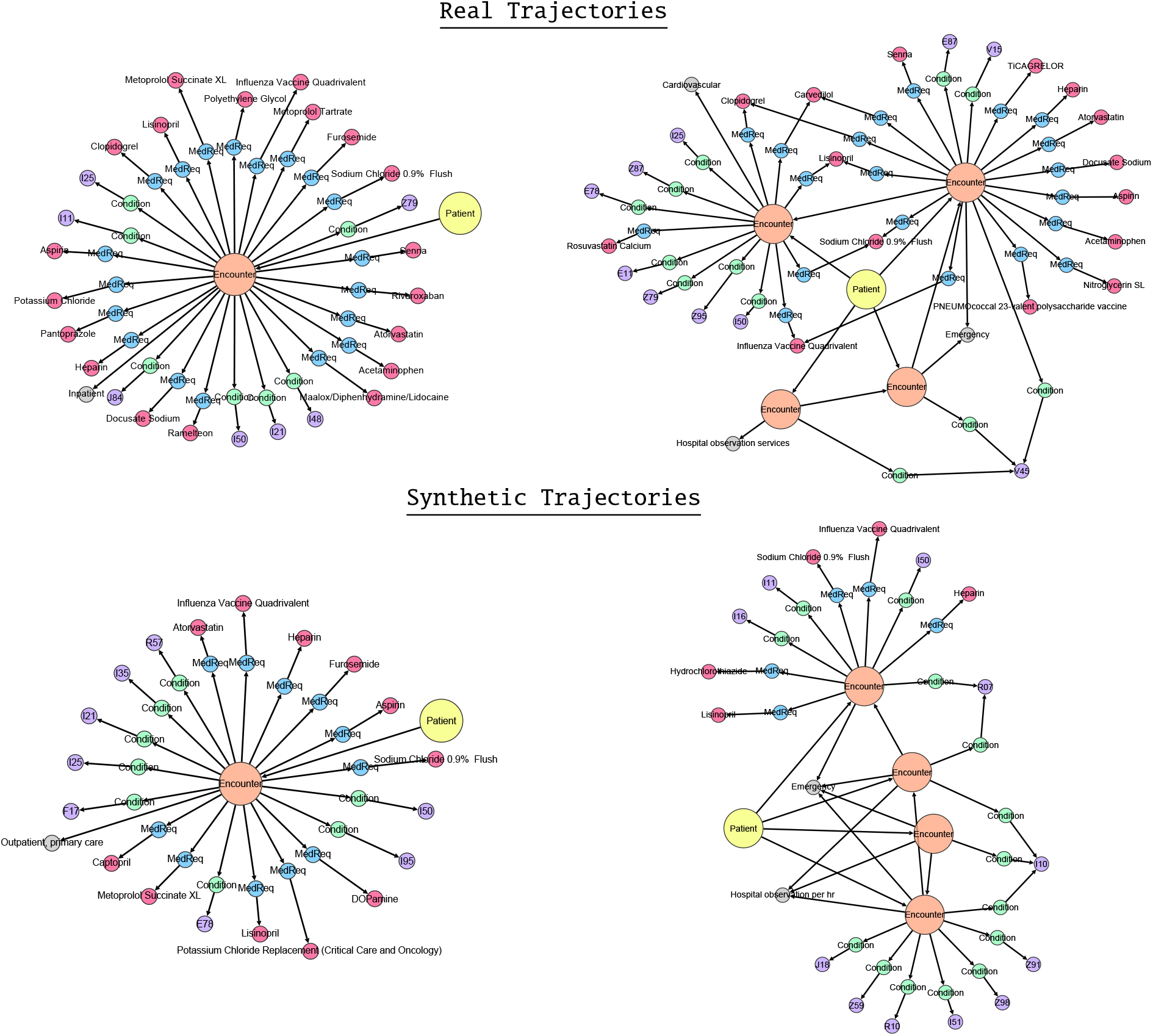
Visualizations of patient trajectories. (a) Two examples of trajectories extracted from the training data. (b) Two examples of synthetic trajectories that were generated with the proposed model.

We also trained a Support Vector Machine (SVM) classifier to learn to distinguish between real trajectories and the synthetic trajectories generated by the model. However, the classifier could only discern that synthetic trajectories were slightly more similar to each other than to the real trajectories. Those results are provided in Table 5 in the Supplementary Materials.

**Table 5:** Classification accuracy on the test data set in the task of distinguishing between real and synthetic patient trajectories. 6535 synthetic trajectories were generated to train a SVM classifier (together with equally many real trajectories) to distinguish between real and synthetic trajectories. Both Weisfeiler-Lehman (WL) and shortest path (SP) graph kernels were used. Two scenarios were investigated. Scenario 1 accounts only for graph structure while scenario 2 in addition considers node labels. The results showed that including node labels increased the classifier’s prediction, indicating that some complex data patterns were more (or less) common in the synthetic data.

|  | Without Labels | With Labels |
| --- | --- | --- |
| WL | 58% | 72% |
| SP | 56% | 73% |

## 3 Discussion

Well generated synthetic healthcare data could provide an opportunity to improve the value of analytics by allowing easier access to data in order to pre-train AI models, generate novel hypotheses, and explore data patterns without jeopardizing patient integrity. In this paper we present a deep learning model for generating synthetic patient trajectories from electronic health records. We show that the model can be effectively trained on real graphs and generate novel ones that are not in the training set. These patient trajectories are clinically realistic while sufficiently different from the trajectories in the training set to preserve patient privacy.

Our model is a Variational Graph Autoencoder (VGAE) tailored to patient trajectories represented as directed acyclic graphs. Previous generative models have failed to produce large graphs or to learn long-range time correlations. The model proposed here solves these issues by decoupling the sequential patient timeline from the clinical interventions. The model is well suited for the complex time-dependencies found in electronic health records. Our numerical results show that the model generates novel synthetic patient trajectories, not found in the training data, that are sufficiently different to preserve patient privacy, yet retains the characteristics of the real-world data. Arguably the most significant feature is that the model is powerful enough to learn long-range correlations between trajectory nodes.

An interesting question rising from this work is to what extent synthetic data can replace real-world data in downstream analysis. Given our experimental results, and the model’s ability to learn paths in patient trajectories, we expect analysis based on either real or synthetic to lead to similar conclusions. This could be tested in practice by comparing output of more machine learning classifiers trained on synthetic trajectories against those trained on real-world data. Are the data-driven insights identical?

As we have already emphasized, the success of any deep learning model rests on the quality and amount of training data. The model can capture general trends already from limited training data, but ultimately requires large amounts of training data to generate long and accurate patient trajectories. In short, as always, the more data the better results. In general, outliers (patient trajectory groups that rarely occur in the training set) are also difficult to generate with accuracy. The generative model should always take measures to ensure that all trajectories of interest are well-represented in the training set.

### Technical limitations

First, node and edge labels is the only metadata included in our model. There is a lot of additional metadata in EHR systems (for example, lab data including values and units) that is of interest for analysis. Such data is represented as key/value pairs on nodes and edges in our graph model. Our generator is easily extended to node and edge attributes by coupling a multi-layer perceptron (MLP) to the model once the node type has been determined. Second, the present version of the model assumes that Encounter nodes are connected to only one type of Condition or Medication node. In practice, there can be more than one on the same node (if, for example, the patient is administered the same medication multiple times in the same encounter). This limitation is due to the model’s binary classifier, which decides whether (or not) a single node of each type should be added to the encounter. A future iteration of the model could replace the binary classifier with a module which accounts for multiplicity. Third, the model has a number of hyper-parameters (see the Methods section) that could be investigated for further sensitivity analysis and optimization.

### Biased synthetic data

Data bias (i. e., skewed representations of certain data categories based on e. g., ethnicity, gender, or something else) in healthcare AI systems has been debated because of its potential to harm misrepresented population groups [50, 51]. Although biased data bias is a real concern to systematic use of healthcare data in general, it is important to separate the discussion regarding bias in the underlying training data from any eventual bias introduced by the AI model itself. We have identified no such model bias.

Synthetic data is not meant to straight-off replace original patient data. Instead, synthetic data can first increase data efficiency by providing a safe and accessible environment for analytics and pre-training AI models. Tentative conclusions from analytics will still need to be verified on original data, as before. But crucially, synthetic data avoids the need to access sensitive real-world data when the chances for meaningful conclusions are lower.

### Privacy risks in the context of synthetic data

A major threat is when a malicious agent can use the synthetic patient trajectories to re-identify real patients from the training data. This is called privacy leaking. That risk is magnified when the agent is in possession of additional information about the real individuals (medical conditions, prescriptions, etc.) that can be combined with the synthetic data to form recognizable patterns that can be used for re-identification. In our model, the amount of similarity between the synthetic and real trajectories is adjustable by the amplitude of the noise injected into the sampled latent space. Synthetic data should undergo a careful evaluation with respect to identity disclosure risks prior to distribution [52]. A number of different approaches for reducing the risk of information disclosure [53, 54] has been proposed, since disclosure control methods have a significant impact on data utility.

### Conclusion and perspectives

Graph deep learning is a powerful tool for learning complex data patterns. Here, a variant of a variational graph autoencoder (VGAE) tailored to patient trajectories represented as large directed acyclic graphs created privacy-preserving and highly accurate synthetic EHRs with long-range time correlations. This approach could reduce the problem of restricted access to health data, thus enabling explorative analyses, algorithm pre-training, hypothesis generation, and data expansion without jeopardizing privacy.

## 4 Methods

### 4.1 Notation

Let [*n*] = {1, …, *n*} ⊂ℕ for *n* ≥1 and *G* = (*V, E*) be a directed graph where *V* is the node set and *E* is the edge set, such that *n* is the number of nodes and *m* is the number of edges in the graph. The neighbourhood *v* of a node *v* is the set of all nodes adjacent to *v*. For a directed graph, we use 𝒩^+ (^*v*) = *u*{|(*v, u*) ∈ *E*} to indicate the set of out-neighbors of *v* where *v, u* is an edge between nodes *v* and *u* of *V*, and 𝒩^− (^*v*) = {*u*| *u, v*) ∈ *E*} to indicate the set of in-neighbors of *v*. The out-degree of node *v* is *d*^+^ (*v*) = 𝒩^+^ (*v*) and its in-degree is *d*^−^ (*v*) = 𝒩^−^ (*v*). The adjacency matrix **A** ∈ ℝ^*n*×*n*^ of a graph *G* is a symmetric (and typically sparse) matrix used to encode edge information in the graph. Element *i, j* is the weight of the edge between nodes *v*_*i*_ and *v* _*j*_ if the edge exists and 0 otherwise. For graphs with node labels and edge labels, nodes and edges are associated with discrete labels, expressed by two functions *ℓ*_*V*_ : *V* →∑_*V*_ and *ℓ*_*E*_ : *E* →∑_*E*_ that map nodes and edges to labels from the sets of labels ∑_*V*_ and ∑_*E*_, respectively.

### 4.2 Architectural details

We designed a model tailored to patient trajectories where each graph corresponds to:

- A Patient node followed by a sequence of Encounter nodes (Figure 11a).
- Each Encounter node is connected to Condition and MedicationRequest nodes, which in turn are terminated with ConditionType and MedicationType nodes. An Encounter node could also be connected to EncounterType and/or EncounterCategory nodes (Figure 11b).

**Figure 11:**
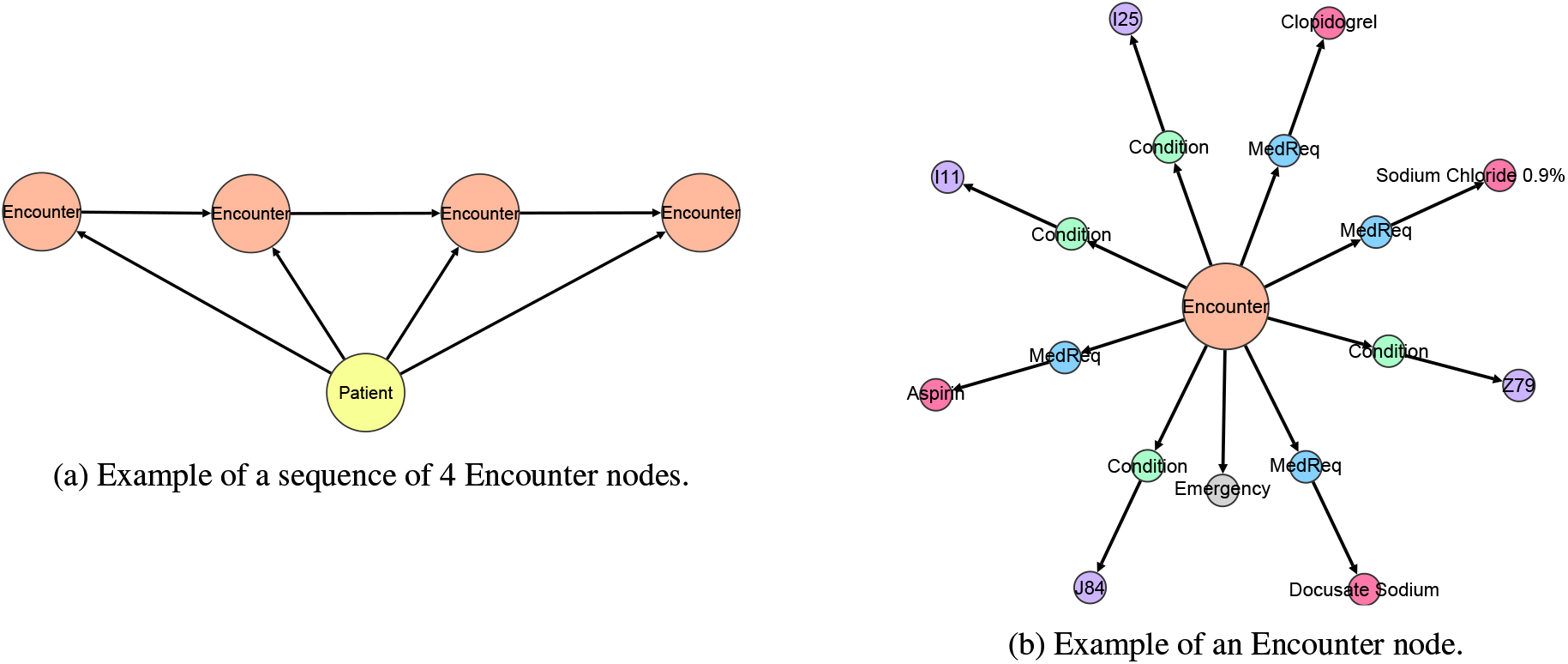
The two main structures into which a patient trajectory can be decomposed. (a) Patient node followed by a sequence of Encounter nodes; and (b) set of Encounter nodes, each connected to several Condition and MedicationRequest nodes, which in turn are terminated with ConditionType and MedicationType nodes.

Clearly, the graph generation can be carried out in two steps: (1) Generate the Encounter node sequence. (2) Generate the successors of each Encounter node. For the first task, we use the topological order of the patient trajectory subgraph obtained only from the Patient and Encounter nodes. This topological order is important because it keeps the trajectory timeline by enforcing Encounter node *u* to precede Encounter *v* chronologically. Since Patient and Encounter nodes are only a small fraction of the nodes in the patient trajectory, a recurrent neural network (RNN) can capture the relationships between consecutive encounters in the sequence. For the second task, we could generate successors of the Encounter nodes by imposing any topological ordering and let another RNN learn that structure. That is possible since Encounter nodes do not have too many successors. In this work, we used an alternative approach where we consider the Encounter successor nodes as a set, and then we simply generate a set that contains those nodes.

We use an encoder-decoder architecture. The encoder maps input DAGs to a distribution parameterized as a multivariate Gaussian. In other words, the encoder predicts the mean and standard deviation of this Gaussian distribution. A random sample is then drawn from the distribution and serves as the latent representation of the input graph. The decoder tries to reconstruct the input DAGs given their vector representations. The decoder is a variational approximation, *p* _*θ*_ (*G*| **z)**, which takes an embedding **z** as input. An overview of the proposed model is given in Figure 12.

**Figure 12:**
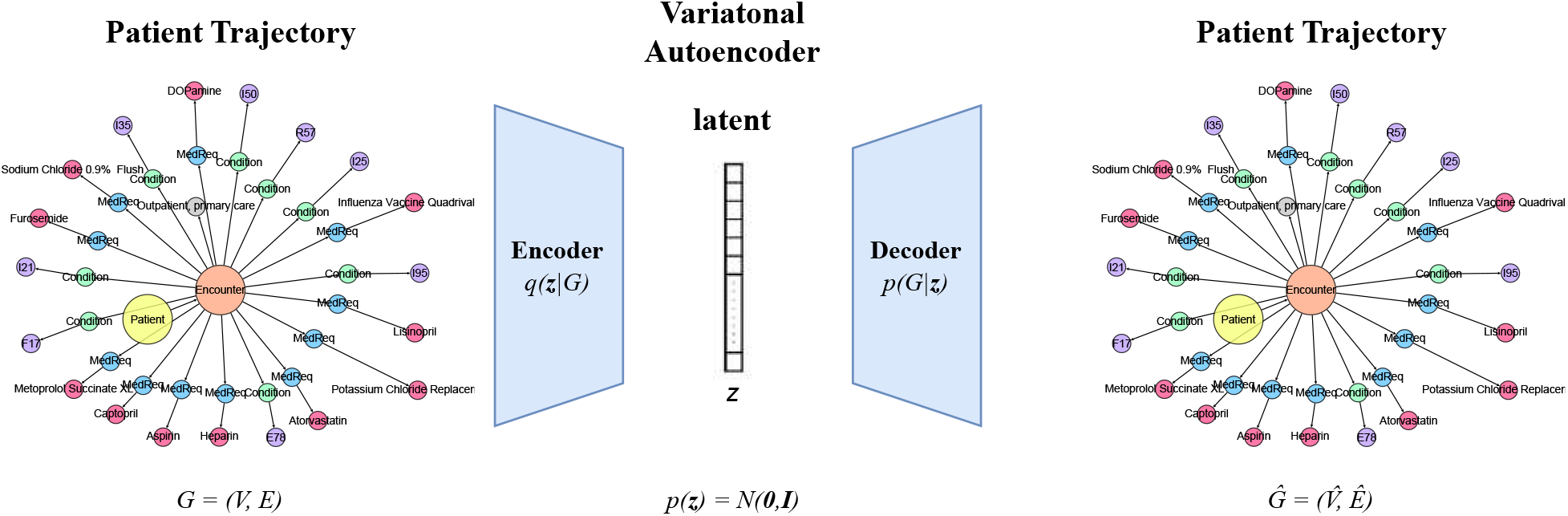
An overview of the proposed variational autoencoder. To train the model, a patient trajectory is fed to the encoder which projects the trajectory to a distribution parameterized as a multivariate Gaussian. Then a latent vector **z** is sampled from that distribution and is passed on to the decoder which reconstructs the input trajectory. To generate a synthetic sample, a vector is sampled from the normal distribution and is then fed directly to the decoder which constructs the synthetic trajectory.

Two pre-processing steps were applied to the patient trajectories before encoding. First, we merged Condition/ConditionType and MedicationRequest/Medication type node pairs. Second, for each graph, an End node was added via a directed edge to the last Encounter node. This allows the model to decide when to terminate the generation of nodes in a new graph.

#### Encoder

The encoder of the model is a message passing graph neural network. Its first part is an embedding layer that creates representations for the nodes in each patient DAG. Each node *v* has a trainable node embedding **x**_*v*_, and there is a single node embedding for each node type. These node embeddings are updated during training with a combination of synchronous and asynchronous message passing schemes.

First, the Encounter node embeddings are updated by aggregating the embeddings of their successors, excluding Encounter and End nodes:

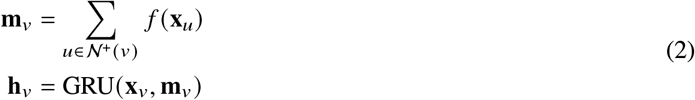

where 𝒩^+ (^*v*) is the set of successors of Encounter node *v* (again, excluding Encounter and End nodes), *f* is a neural network (MLP), **x**_*v*_ is the embedding of node *v*, and GRU is a gated recurrent unit.

An asynchronous message passing scheme is then applied where we sequentially perform message passing according to the topological sorting obtained from the patient subgraph of Encounter and End nodes. This differs from the standard message passing scheme in graph neural networks where all node embeddings are updated at each algorithm step. In our algorithm, the node embeddings are updated in this step according to:

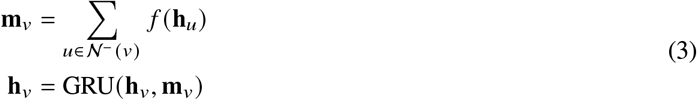

where 𝒩^−^ (*v*) is the set of incoming neighbors of *v* for Encounter nodes.

Once all node embeddings of the DAG have been computed, we use the end node embedding (i. e., the node without any successors) as the output of the encoder. Thus, **h**_*G*_ = **h**_*e*_ where *e* denotes the End node of *G*. This vector is passed to two fully-connected layers to get the mean and variance parameters of the posterior approximation *q* (**z**|*G*):

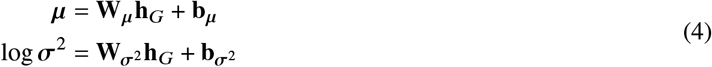

#### Decoder

The decoder of the model also applies an asynchronous message passing scheme to generate node representations. The decoder uses a GRU to update node embeddings when generating the graph.

A fully-connected layer is used to map the input latent vector **z** to the initial (hidden) state vector **h**_0_. The state vector is passed to the GRU, which constructs a DAG node-by-node. So far, all are Encounter (or End) nodes. The embedding of the first (Patient) node is 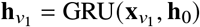. The following steps are performed to generate node *v*_*i*_:

1. Compute the label distribution of *v*_*i*_ with an MLP based on the current graph state 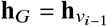.
2. Sample the label of *v*_*i*_. If this is the end label, stop the decoding, connect the last Encounter node to *v*_*i*_, and return the DAG. If not, continue the generation.
3. Connect the last added Encounter node and the Patient node to *v*_*i*_. Update **h**_*vi*_ according to:
4. Produce a vector **s** ∈**ℝ**^*c*^ (*c* denotes the different types of successors of Encounter nodes excluding Encounter and End nodes):

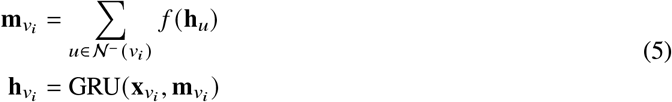

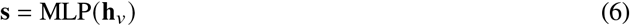

The sigmoid function is applied point-wise to the MLP output and then the model decides whether to add a node of each type of successor to the graph. When a new node is added, so is a directed edge from *v*_*i*_. The decision to add a successor to the graph is a binary classification problem. We therefore use the binary cross entropy loss to train the model.

### 4.3 Loss function

The loss function of our variational autoencoder has two terms,

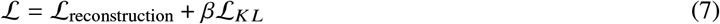

The first term is the reconstruction loss, i. e., the variational lower bound, and measures how well the model reconstructs the input data. The reconstruction loss is high if the reconstructed DAG is very different from its input. This term can be split into two contributions, ℒ_reconstruction_ = ℒ_encounter_ + ℒ_other_. One contribution measures how well the model can reconstruct the sequence of Encounter nodes. It is equal to the binary cross-entropy between the predicted types of Encounter or End nodes and their actual types:

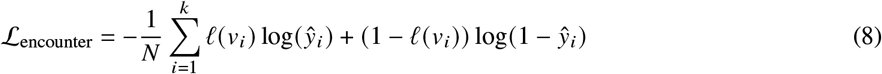

Remember, {*v*_1_, …, *v*_*k*_} are only the Encounter and End nodes in the DAG. The other contribution measures how well the model can reconstruct the successors of the Encounter nodes. It is equal to the binary cross-entropy between the predicted and the actual successors of each Encounter node:

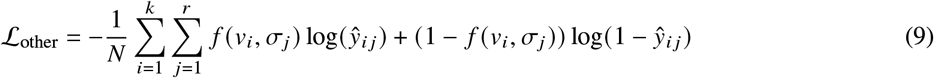

Here the {*σ*_1_, …, *σ*_*r*_} set includes all nodes except Encounter and End nodes for node *v*:

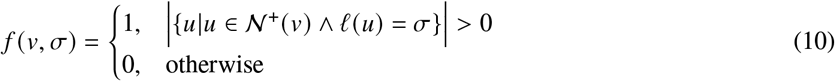

The second term of the loss function is a regularization term. It is equal to the Kullback–Leibler (KL) divergence of the approximate *q* (**z**|*G*) from the true posterior *p* (**z**), where *p* (**z**) = 𝒩 (**0, I**) and **0** and **I** are the all-zeros vector and the identity matrix, respectively. The KL divergence measures how closely the output distribution *q* (**z**|*G*) matches *p* (**z**):

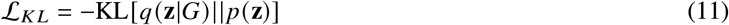

### 4.4 Training

To train the model, we used teacher forcing [55], i. e., a strategy for training RNNs that feeds the observed sequence values as input to the model, instead of feeding the model’s output from a prior time step, thus forcing the model to stay close to the ground-truth sequence. Given the topological ordering of the Encounter nodes, in each decoding step, we force the model to generate the ground-truth node type (Encounter node or End node) and the ground-truth condition and medication nodes connected to the Encounter nodes. Note that during generation of new samples, teacher forcing cannot be applied since there is no ground-truth information, and thus we sample node types according to the decoding distributions. We used a cyclical scheme for the annealing parameter in equation (7) which increases *β* multiple times [56]. This is done to balance the two terms in the loss function such that the model learns the underlying distribution while maximizing the use of the avaiable latent space.

### 4.5 Experimental Setup

We used the following values for the model’s hyper-parameters. The hidden-dimension size of the embedding layer and the GRU layers were 512. The hidden-dimension size of the fully-connected layer that transforms the sampled vector representation of the graphs was set to 512 and followed by a tanh-activation. We used an MLP with hidden-dimension size 1024 to decide whether a new node type was to be added to the graph (and also to determine its type). We used an MLP with hidden-dimension size 2048 to compute the successors of Encounter nodes. The hidden layers in both MLPs were followed by ReLU activation functions. The dimension of the multivariate Gaussian distribution was set to 256. The batch size was 256 and the number of learning epochs was 5000. We used the Adam optimizer with an initial learning rate of 10^−3^ and decayed the learning rate by 0.1 every 1000 epochs to a minimum of 10^−5^. The model with the lowest training loss was stored on disk and retrieved at the end of training. The best model was then used to generate new graphs for the numerical experiments.

## Data Availability

All data produced are available online at PhysioNet repository (https://physionet.org). Access is authorized to users through a data use agreement with the providers.

## Data Availability

MIMIC-IV data is available on the PhysioNet repository (https://physionet.org/) and access is authorized to users through a data use agreement with the providers.

## Code Availability

The code of the presented model together with the code for the statistical analyses is available upon request to the corresponding author.

## Acknowledgements

M.V. is partially supported by the “Wallenberg AI, Autonomous Systems and Software Program” (WASP). M.L. is partially supported by AIR Lund (Artificially Intelligent use of Registers at Lund University) research environment, and received funding from the Swedish Research Council (VR; grant no. 2019-00198). G.N. is supported by the French National research agency via the AML-HELAS (ANR-19-CHIA-0020) project.

## Author Contributions

E.G.B. designed and directed this study. G.N. and M.V. contributed to methodology design. G.N. conducted all the experiments. G.N., M.V., M.L. and E.G.B. analyzed and discussed the results. All authors wrote, reviewed, and revised the manuscript.

## Supplementary Materials

## References

[1] N. Rieke, et al., The future of digital health with federated learning. npj Digital Medicine 3(1), 1–7 (2020)

[2] M. Abadi, et al., in Proceedings of the 2016 ACM SIGSAC Conference on Computer and Communications Security (2016), pp. 308–318

[3] A. Acar, H. Aksu, A.S. Uluagac, M. Conti, A survey on homomorphic encryption schemes: Theory and implementation. ACM Computing Surveys 51(4), 1–35 (2018)

[4] J. Yoon, D. Jarrett, M. Van der Schaar, in Advances in Neural Information Processing Systems (2019)

[5] G. Ramponi, P. Protopapas, M. Brambilla, R. Janssen, T-cgan: Conditional generative adversarial network for data augmentation in noisy time series with irregular sampling. arXiv preprint arXiv:1811.08295 (2018)

[6] S. Kuutti, R. Bowden, Y. Jin, P. Barber, S. Fallah, A survey of deep learning applications to autonomous vehicle control. IEEE Transactions on Intelligent Transportation Systems 22(2), 712–733 (2020)

[7] M. Popel, et al., Transforming machine translation: a deep learning system reaches news translation quality comparable to human professionals. Nature communications 11(1), 1–15 (2020)

[8] W.P. Walters, R. Barzilay, Applications of deep learning in molecule generation and molecular property prediction. Accounts of chemical research 54(2), 263–270 (2020)

[9] E. Choi, et al., in Proceedings of Machine Learning for Healthcare 2017 (2017), pp. 286–305

[10] J. Jordon, J. Yoon, M. Van Der Schaar, in 7th International Conference on Learning Representations (2019)

[11] C. Esteban, S.L. Hyland, G. Rätsch, Real-valued (medical) time series generation with recurrent conditional gans. arXiv preprint arXiv:1706.02633 (2017)

[12] P. Wendland, et al., Generation of realistic synthetic data using multimodal neural ordinary differential equations. npj Digital Medicine 5(1), 1–10 (2022)

[13] A. Tucker, Z. Wang, Y. Rotalinti, P. Myles, Generating high-fidelity synthetic patient data for assessing machine learning healthcare software. npj Digital Medicine 3(1), 1–13 (2020)

[14] R.J. Chen, M.Y. Lu, T.Y. Chen, D.F. Williamson, F. Mahmood, Synthetic data in machine learning for medicine and healthcare. Nature Biomedical Engineering 5(6), 493–497 (2021)

[15] A. Goncalves, et al., Generation and evaluation of synthetic patient data. BMC medical research methodology 20(1), 1–40 (2020)

[16] D.P. Kingma, M. Welling, in 2nd International Conference on Learning Representations (2014)

[17] D.P. Kingma, M. Welling, An introduction to variational autoencoders. Foundations and Trends(r) in Machine Learning 12(4), 307–392 (2019)

[18] M. Simonovsky, N. Komodakis, in Proceedings of the 27th International Conference on Artificial Neural Networks (2018), pp. 412–422

[19] G. Salha, S. Limnios, R. Hennequin, V.A. Tran, M. Vazirgiannis, in Proceedings of the 28th ACM International Conference on Information and Knowledge Management (2019), pp. 589–598

[20] M. Chatzianastasis, G. Dasoulas, G. Siolas, M. Vazirgiannis, in Proceedings of the 2021 IEEE/CVF International Conference on Computer Vision Workshops (2021), pp. 393–402

[21] A. Creswell, et al., Generative adversarial networks: An overview. IEEE Signal Processing Magazine 35(1), 53–65 (2018)

[22] J. Gui, Z. Sun, Y. Wen, D. Tao, J. Ye, A review on generative adversarial networks: Algorithms, theory, and applications. IEEE Transactions on Knowledge and Data Engineering (2021)

[23] D. Kaur, et al., Application of bayesian networks to generate synthetic health data. Journal of the American Medical Informatics Association 28(4), 801–811 (2021)

[24] J. Walonoski, et al., Synthea: An approach, method, and software mechanism for generating synthetic patients and the synthetic electronic health care record. Journal of the American Medical Informatics Association 25(3), 230–238 (2018)

[25] M.K. Baowaly, C.C. Lin, C.L. Liu, K.T. Chen, Synthesizing electronic health records using improved generative adversarial networks. Journal of the American Medical Informatics Association 26(3), 228–241 (2019)

[26] A. Yale, et al., Generation and evaluation of privacy preserving synthetic health data. Neurocomputing 416, 244–255 (2020)

[27] T.N. Arvanitis, S. White, S. Harrison, R. Chaplin, G. Despotou, A method for machine learning generation of realistic synthetic datasets for validating healthcare applications. Health Informatics Journal 28(2) (2022)

[28] K. Chin-Cheong, T. Sutter, J.E. Vogt, in Workshop on Machine Learning for Health (ML4H) at the 33rd Conference on Neural Information Processing Systems (2019)

[29] D. Saxena, J. Cao, Generative adversarial networks (gans) challenges, solutions, and future directions. ACM Computing Surveys 54(3), 1–42 (2021)

[30] J. You, R. Ying, X. Ren, W. Hamilton, J. Leskovec, in Proceedings of the 35th International Conference on Machine Learning (2018), pp. 5708–5717

[31] W. Jin, R. Barzilay, T. Jaakkola, in Proceedings of the 35th International Conference on Machine Learning (2018), pp. 2323–2332

[32] Y. Li, O. Vinyals, C. Dyer, R. Pascanu, P. Battaglia, in Proceedings of the 35th International Conference on Machine Learning (2018)

[33] P. Bongini, M. Bianchini, F. Scarselli, Molecular generative graph neural networks for drug discovery. Neurocomputing 450, 242–252 (2021)

[34] A. Johnson, et al. Mimic-iv (2021). URL https://physionet.org/content/mimiciv/1.0/

[35] Implemented in the SHAARPEC Analytics platform.:https://www.shaarpec.com.

[36] D. Bender, K. Sartipi, in Proceedings of the 26th IEEE International Symposium on Computer-Based Medical Systems (2013), pp. 326–331

[37] E. Jang, S. Gu, B. Poole, in 5th International Conference on Learning Representations (2017)

[38] N. De Cao, T. Kipf, Molgan: An implicit generative model for small molecular graphs. arXiv preprint arXiv:1805.11973 (2018)

[39] G. Nikolentzos, G. Siglidis, M. Vazirgiannis, Graph kernels: A survey. Journal of Artificial Intelligence Research 72, 943–1027 (2021)

[40] N. Shervashidze, P. Schweitzer, E.J. Van Leeuwen, K. Mehlhorn, K.M. Borgwardt, Weisfeiler-lehman graph kernels. Journal of Machine Learning Research 12(9) (2011)

[41] K.M. Borgwardt, H.P. Kriegel, in Proceedings of the 5th IEEE International Conference on Data Mining (2005)

[42] B. Weggenmann, V. Rublack, M. Andrejczuk, J. Mattern, F. Kerschbaum, in Proceedings of the ACM Web Conference 2022 (2022), pp. 721–731

[43] W. Kawai, Y. Mukuta, T. Harada, Scalable generative models for graphs with graph attention mechanism. arXiv preprint arXiv:1906.01861 (2019)

[44] A. Gretton, K.M. Borgwardt, M.J. Rasch, B. Schölkopf, A. Smola, A Kernel Two-Sample Test. The Journal of Machine Learning Research 13(1), 723–773 (2012)

[45] J. Engdahl, A. Holmén, M. Rosenqvist, U. Strömberg, Uptake of atrial fibrillation screening aiming at stroke prevention: geo-mapping of target population and non-participation. BMC Public Health 13(1), 715–724 (2013)

[46] W.G. Members, V.L. Roger, A.S. Go, D.M. Lloyd-Jones, E.J. Benjamin, J.D. Berry, W.B. Borden, D.M. Bravata, S. Dai, E.S. Ford, et al., Heart disease and stroke statistics—2012 update: a report from the american heart association. Circulation 125(1), e2.–e220 (2012)

[47] T. Vos, et al., Global, regional, and national incidence, prevalence, and years lived with disability for 310 diseases and injuries, 1990–2015: a systematic analysis for the global burden of disease study 2015. The Lancet 388(10053), 1545–1602 (2016)

[48] B.J. Mortazavi, et al., Analysis of machine learning techniques for heart failure readmissions. Circulation: Cardiovascular Quality and Outcomes 9(6), 629–640 (2016)

[49] B. Schölkopf, J.C. Platt, J. Shawe-Taylor, A.J. Smola, R.C. Williamson, Estimating the support of a highdimensional distribution. Neural Computation 13(7), 1443–1471 (2001)

[50] N. Mehrabi, F. Morstatter, N. Saxena, K. Lerman, A. Galstyan, A survey on bias and fairness in machine learning. ACM Comput. Surv. 54(6), 1–35 (2021)

[51] R.B. Parikh, S. Teeple, A.S. Navathe, Addressing bias in artificial intelligence in health care. JAMA 322(24), 2377–2378 (2019)

[52] J.P. Reiter, R. Mitra, Estimating risks of identification disclosure in partially synthetic data. Journal of Privacy and Confidentiality 1(1) (2009)

[53] J.P. Reiter, Satisfying disclosure restrictions with synthetic data sets. Journal of Official Statistics 18(4), 531 (2002)

[54] N. Park, et al., Data synthesis based on generative adversarial networks. Proceedings of the VLDB Endowment 11(10) (2018)

[55] R.J. Williams, D. Zipser, A learning algorithm for continually running fully recurrent neural networks. Neural Computation 1(2), 270–280 (1989)

[56] H. Fu, et al., in Proceedings of the 2019 Conference of the North American Chapter of the Association for Computational Linguistics: Human Language Technologies (2019), pp. 240–250

